# A targeted behavioural economics-based intervention to improve medication adherence in patients with Hypertension: the BEHAVE-HTN study protocol

**DOI:** 10.64898/2025.12.12.25342134

**Authors:** Annina S. Vischer, Jan Strebel, Anjali Raja Beharelle, Michael Mayr, Valeriya Nemtsova, Gregoire Wuerzner, Georg Ehret, Michael Stiefel, Emrush Rexhaj, Théo Meister, Thilo Burkard

## Abstract

**Background and Aims:** Non-adherence to antihypertensive therapy is a major contributor to poor blood pressure (BP) control, increased cardiovascular risk, and higher healthcare costs. Behavioral economics suggests that financial incentives and mobile-based interventions may support behavior change, yet evidence for such strategies in central Europe is limited.

**Objective:** The BEHAVE-HTN study evaluates whether a multi-faceted medication adherence mobile application (mApp) with behavioral economic principles—including reminders, gamification, and financial incentives—improves medication adherence in patients with hypertension compared to standard of care.

**Methods:** This multi-center, randomized, open-label trial will include 180 outpatients with hypertension taking four or more pills daily. Participants will be randomized into three groups (n = 60 per group): (A) mApp with contingent financial incentives, (B) mApp without incentives, and (C) standard of care. All participants will receive a Medication Event Monitoring System (MEMS) dispenser to objectively track adherence. The primary endpoint is the difference in mean MEMS adherence between group A and the control group at 90 days. A co-primary endpoint compares MEMS adherence in the final vs. initial month of the intervention. Secondary outcomes include changes in BP, app-reported adherence, and patient-reported feedback on the mApp.

**Results:** Currently, participant inclusion has been nearly completed. We are undergoing data clearing and are planning data analysis. These steps are planned for autumn 2025.

**Conclusion:** This study will provide evidence on whether a multi-faceted medication adherence mobile App — with or without financial incentives—can improve medication adherence in hypertensive patients. The findings may support the implementation of mobile health strategies in routine hypertension care.

**Trial Registration:** SNCTP000004345; ClinicalTrials.gov Identifier: NCT04708756

## Introduction

According the WHO (World Health Organization) global report on hypertension, published in 2023, there is an emerging global burden in hypertension, resulting in cardiovascular disease and all-cause mortality [1]. This report emphasizes the urgent need to address hypertension, its control, and its associated complications, affecting over one billion people worldwide. Non-adherence to antihypertensive therapy is one of the well-recognized major barriers to managing hypertension and to achieving BP control[1,2]. Therefore the WHO stated as early as 2003 that “increasing the effectiveness of adherence interventions may have a far greater impact on the health of the population than any other improvement in specific medical treatments”[3].

Adherence to a medication regimen is defined as “the extent to which patients take their medications as prescribed by their healthcare providers” [4]. Patients are considered adherent if their medication adherence percentage—pills taken divided by pills prescribed within a given period—exceeds 80% [4]. Non-adherence includes failure to initiate therapy, early discontinuation, or incorrect dosage/frequency.

About 50% of patients with chronic illnesses do not take medications as prescribed [3,5], even after a cardiovascular event [6]. Studies show 50–80% non-adherence in hypertensive patients [7], and in Italy, 36% of newly treated patients failed to renew their initial antihypertensive prescription [8].

Medication non-adherence contributes to premature deaths and higher healthcare use. In the U.S., it causes 125,000 deaths and costs USD 100–300 billion annually [9,10]. In Europe, non-adherence is linked to 200,000 premature deaths and EUR 125 billion in excess costs each year [11,12]. Despite numerous interventions, adherence rates have not significantly improved [13,14]. To improve medication adherence, it is important to understand the multifactorial causes of reduced adherence. The WHO classified these factors into 5 categories: socioeconomic factors, factors associated with the health care team and system in place, disease-related factors, therapy-related factors, and patient-related factors [3]. In particular, for therapy-related factors, patients concurrently prescribed multiple medications or who have to take several pills per day, tend to have lower medication adherence [15].

Behavioral interventions, such as reminder systems, pill boxes, and tailored regimens, have shown mixed results in improving medication adherence [16,17]. A review of randomized control trials that seek to improve medication adherence revealed that those that were most effective were multi-faceted, and included combinations of convenience, education, reminders, and reinforcement[18].

To improve the efficacy of these interventions, it’s important to consider deeper behavioral drivers like decision errors. One such error, present bias, leads persons to favor immediate rewards over larger future ones, and has been shown to be a leading cause of non-adherence, particularly in chronic conditions like hypertension where symptoms are absent [5]. One strategy to overcome the present bias is to provide a short-term reward for increased medication adherence, as these external rewards can provide motivation to supplement insufficient intrinsic motivation for behaviors that do not have immediate benefits. A review of 11 U.S. studies found that financial incentives, such as cash, gifts, or vouchers, improved adherence in most cases, however, evidence on the effectiveness of these strategies in Central Europe remains limited [19].

To date, there are no specific approved digital tools recommended for medication adherence in patients with hypertension [20]. The BEHAVE-HTN study therefore intends to test a novel multi-faceted approach to improve medication adherence among patients with hypertension in Switzerland comparing a multi-faceted medication adherence mobile application (mApp) with financial incentives and the mApp without financial incentives against standard of care.

## Methods

### Study rationale

Traditional interventions aimed at improving medication adherence are complex and often ineffective. While a meta-analysis of fourteen studies demonstrates that mobile applications can significantly enhance medication adherence, there is considerable heterogeneity among studies including in patients with hypertension [21,22]. The BEHAVE-HTN study intends to test a novel approach to improving medication adherence. Participants in the intervention groups will participate in a 90-day mApp-based adherence intervention (with or without incentives) based on modern behavioral economics principles plus a 90-day follow-up phase. The intervention will be administered through the Collabree mApp (Collabree AG, Dreikönigstrasse 34, 8002 Zürich, Switzerland).Subjects will be reminded to take their medications based on their digitized medication plan, and they will need to report their daily medication adherence. Participants in the intervention group A will be offered a contingent financial support for app-reported adherence, which consist of clicking a confirmation button in the mApp to report the medication intake at the time of administration. Subjects will have a deduction from total rewards for each non-adherent day, while participants in the intervention group B will receive the mApp without contingent rewards. Avoiding reward loss has been shown to be more motivating than working for reward gain [23]. Additionally, participants in the intervention groups as well as the control group will receive aMEMS dispenser that will record the day and time of dispenser openings.

Using a multi-faceted medication adherence mobile App including contingent rewards, the BEHAVE-HTN study will be able to shed light on the potential of this approach to improve adherence in patients with hypertension, which may result in improved BP control.

### Hypothesis

Medication reminders in connection with contingent rewards administered through the mApp will be superior to standard of care for improving medication adherence in participants with hypertension.

### Multi-faceted medication adherence mApp versus standard of care

The BEHAVE-HTN intervention is a mApp including a multi-faceted approach to target cognitive bias including reminders, contingent rewards for medication adherence, reinforcement for positive behaviors through messages and animations, gamification aspects, and health information provided in a patient friendly form.

By sending timely cues, monitoring the desired action, and rewarding positive behavior in a personalized way, the mApp provides key elements for habit formation in order to support users in establishing healthy and sticky habits and developing goal-directed behaviors and routines [24,25]. In the mApp, patients can easily store their personal medication and therapy plan. Patients can add the desired time and the intervals for taking the medication. After successfully saving the plan, the mApp will remind the patient that it is time to take the medication within a two-hour time window through a push notification. Additionally for one of the intervention groups, it is planned to provide a contingent reward with a deduction for each non-adherent day. The study therefore includes three treatment arms:

#### Group A

Participants in this group receive the mApp intervention with financial incentives. They have the opportunity to keep a maximum of CHF 100 at three time points: after 30 days, 60 days, and 90 days (Endpoint (EP) visit), which. For each missed intake outside the 2-hour window defined at baseline (BL), CHF 4 is deducted per day. Therefore, total earnings range from CHF 0 to CHF 300.

#### Group B

Participants receive the mApp without financial incentives. Both group A and B use the mApp from the BLvisit (day 0) until the extended Follow-up visit (eFU. day 180), after which it is uninstalled.

#### Control group

Participants receive standard care without the mApp or financial incentives.

### Study design

This study has a confirmatory, external, block randomized, stratified, controlled, parallel and open-label design. 180 patients with hypertension will be randomly allocated to either the control group or to the intervention groups (60 patients in each group). We estimate a 10% withdrawal rate based on a similar study [26]. It is intended that dropouts be replaced in a 1:1 fashion as long as the number of withdrawals stays below the estimate of 10% withdrawals. The study will assess the use of a multi-faceted medication adherence mApp with a contingent reward intervention (group A) against the mApp without contingent rewards (group B) and against standard of care (control group) in a representative population of patients with hypertension. Additionally, participants in both the intervention groups as well as the control group will receive a MEMS dispenser that records the day and time of dispenser openings. Outcomes will be assessed using objective data from four in-person visits (0, 45, 90 and 180 days) and the data recorded in the mApp. The data will be collected and compared between groups (Figure 1).

**Figure 1.**
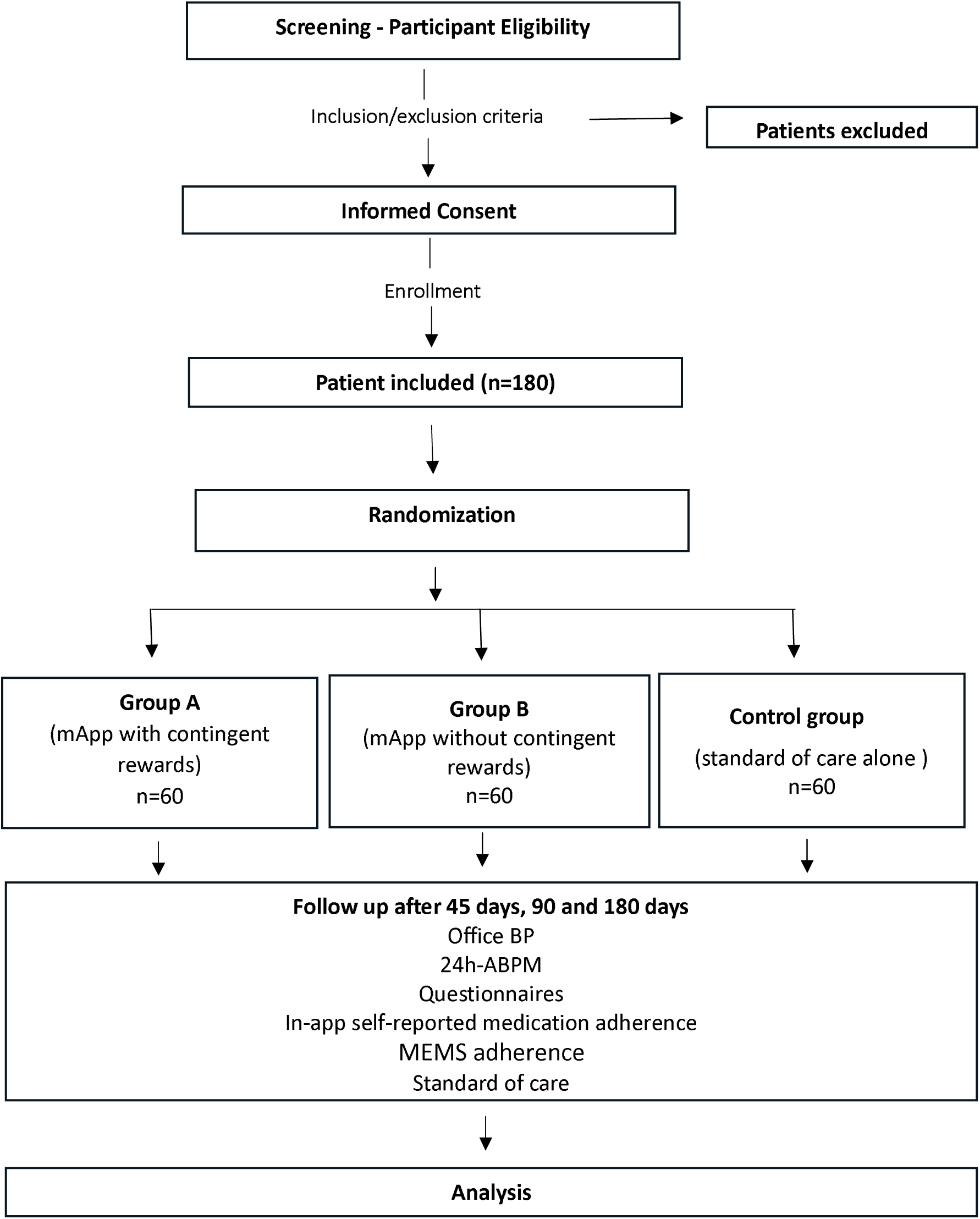
Flow chart of the BEHAVE-HTN clinical trial Abbreviations: BP, Blood Pressure; 24h-ABPM, 24-hour Ambulatory Blood Pressure Monitoring; mApp, multi-faceted medication adherence mobile App; MEMS, Medication Event Monitoring System.

### Recruitment and population

Enrolled patients are required to be diagnosed with hypertension, older than 18 years of age, prescribed 4 or more pills per day, and to meet the predefined inclusion and exclusion criteria to be eligible to participate in this study (Table 1). Potential candidates will be screened for these criteria at the centers. If the criteria are met, the patient will be approached personally to provide written informed consent.

**Table 1.**
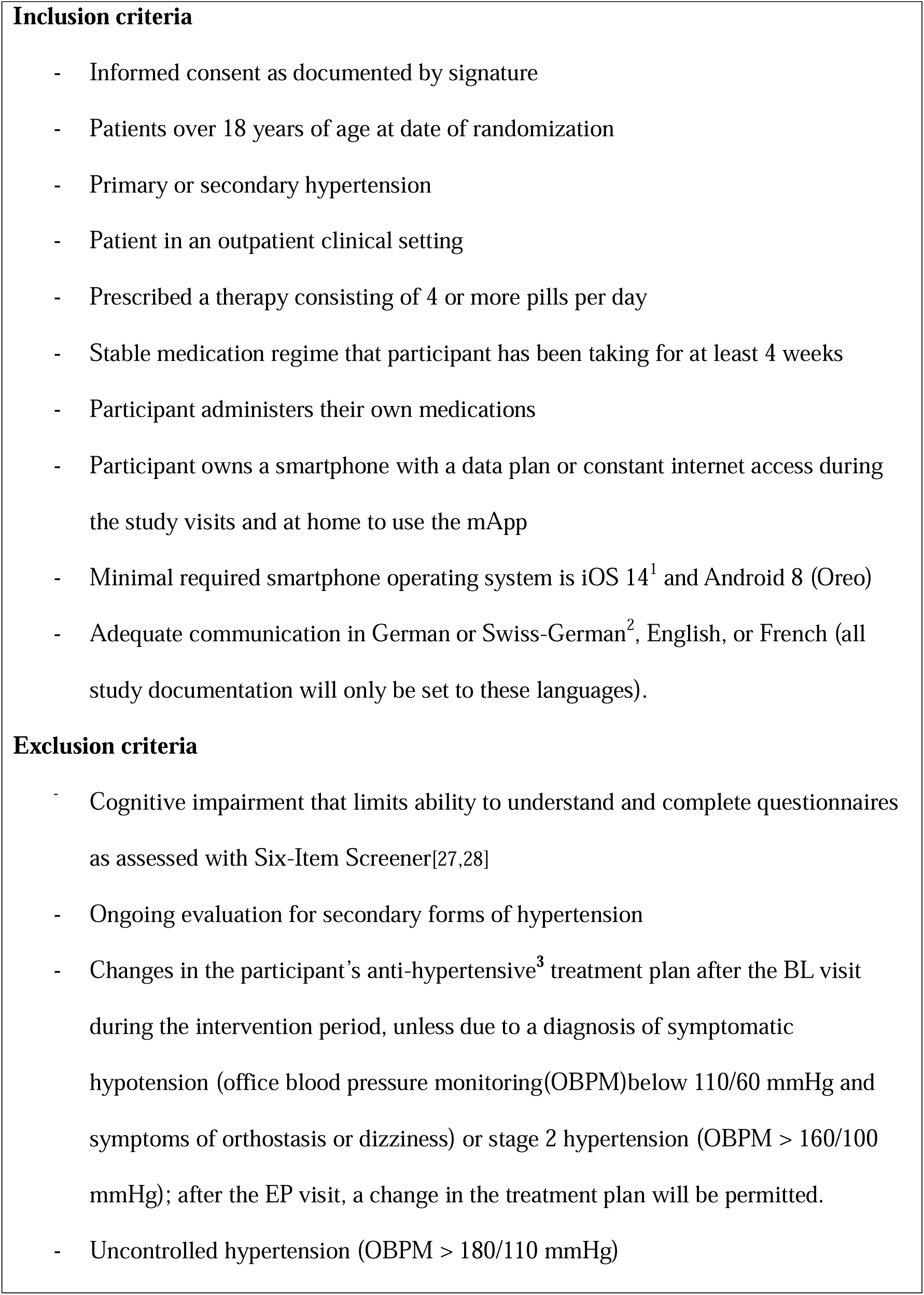
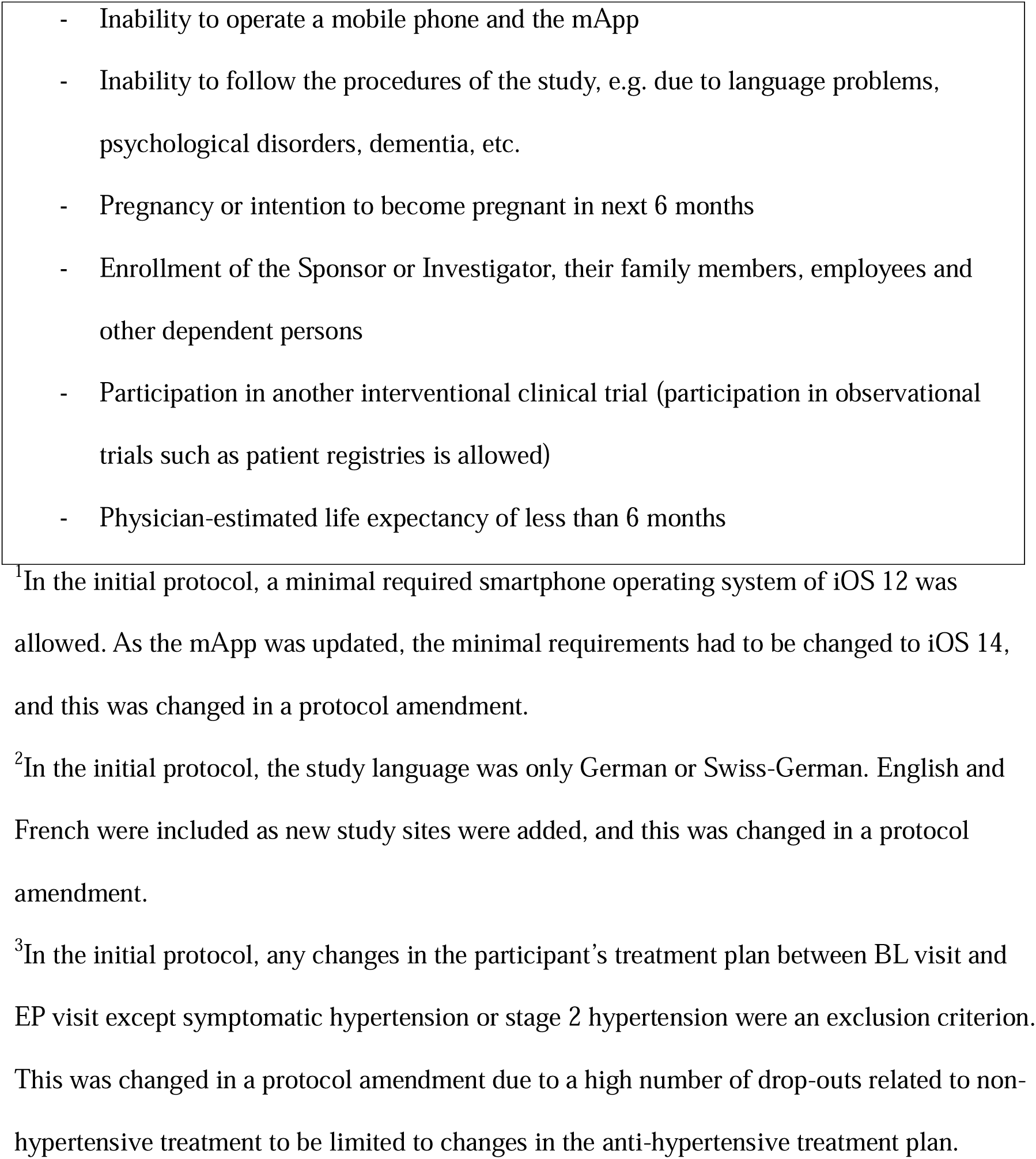
Inclusion and exclusion criteria for the BEHAVE-HTN study.

The investigators will explain to each participant the nature of the study, its purpose, the procedures involved, the expected duration, the potential risks and benefits, and any discomfort it may cause. Additionally, during this initial visit before the randomization of each patient, the physician will conduct a brief discussion of hypertension and the importance of medication adherence. Afterwards, patient data will be entered in the eCRF (electronic Case Report Form), and the patients will be assigned to one of the three study groups. After enrollment, patients will be visited by the study staff, and, once all pre-specified procedures will be executed according to the study plan (Table 2).

**Table 2.**
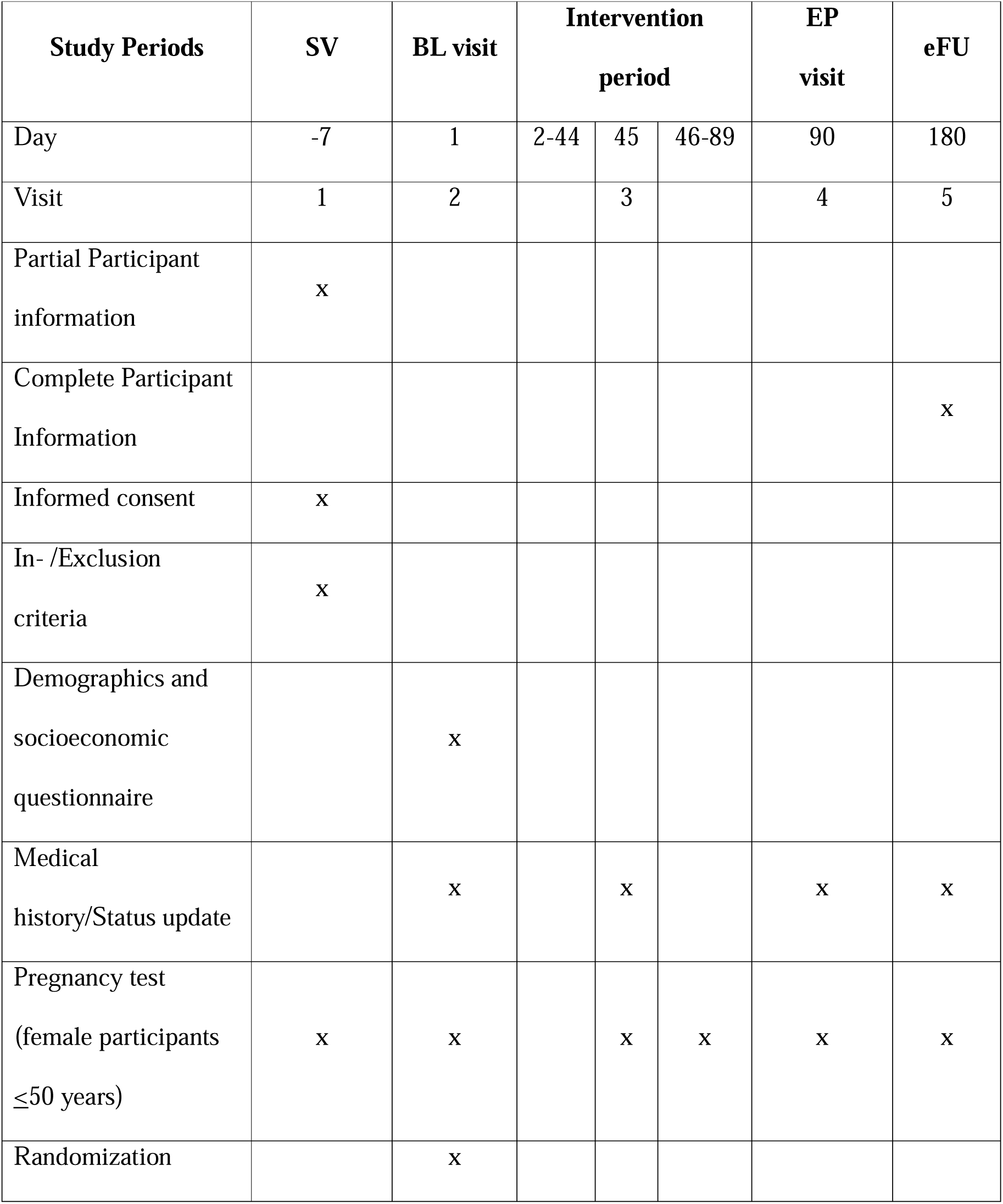
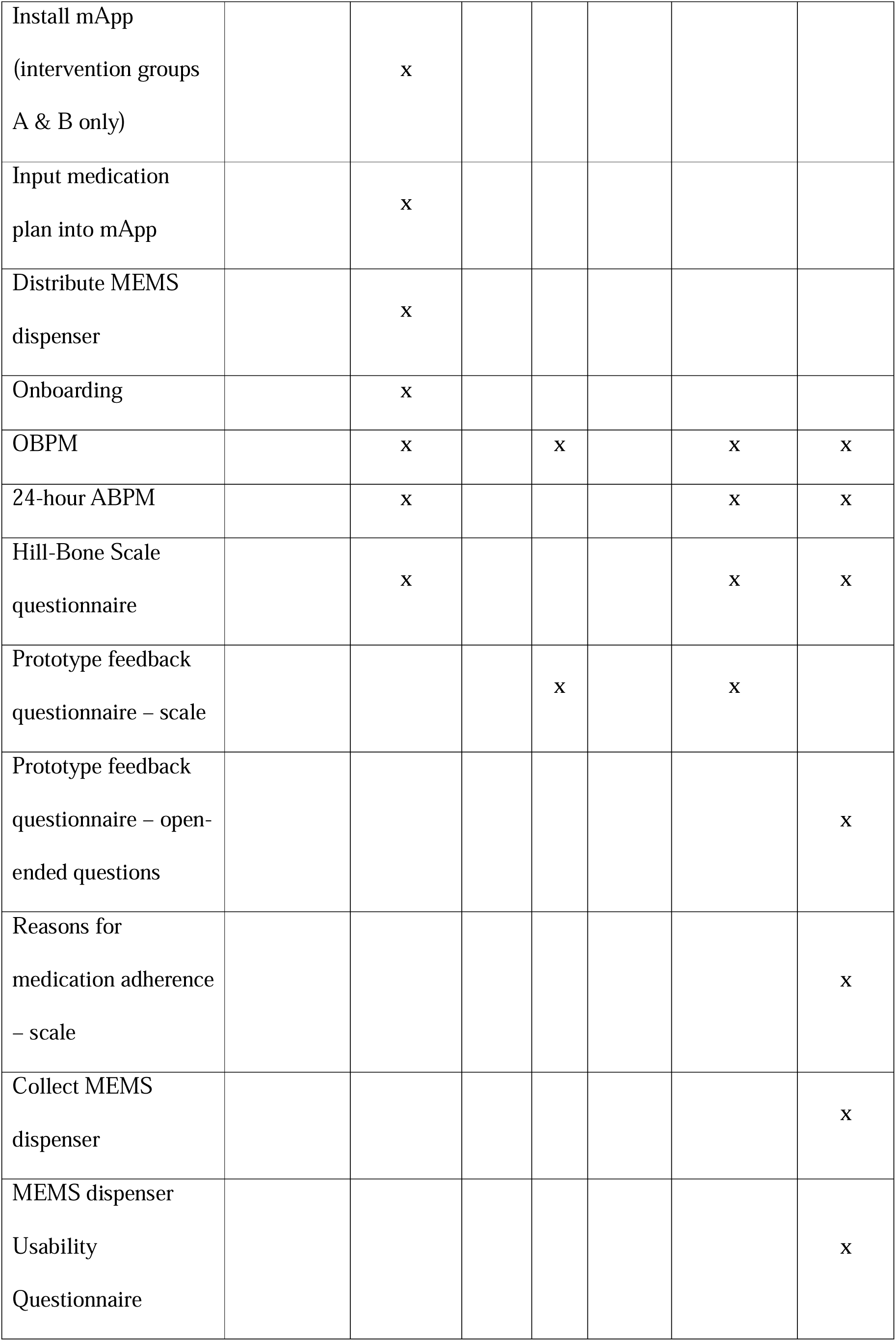
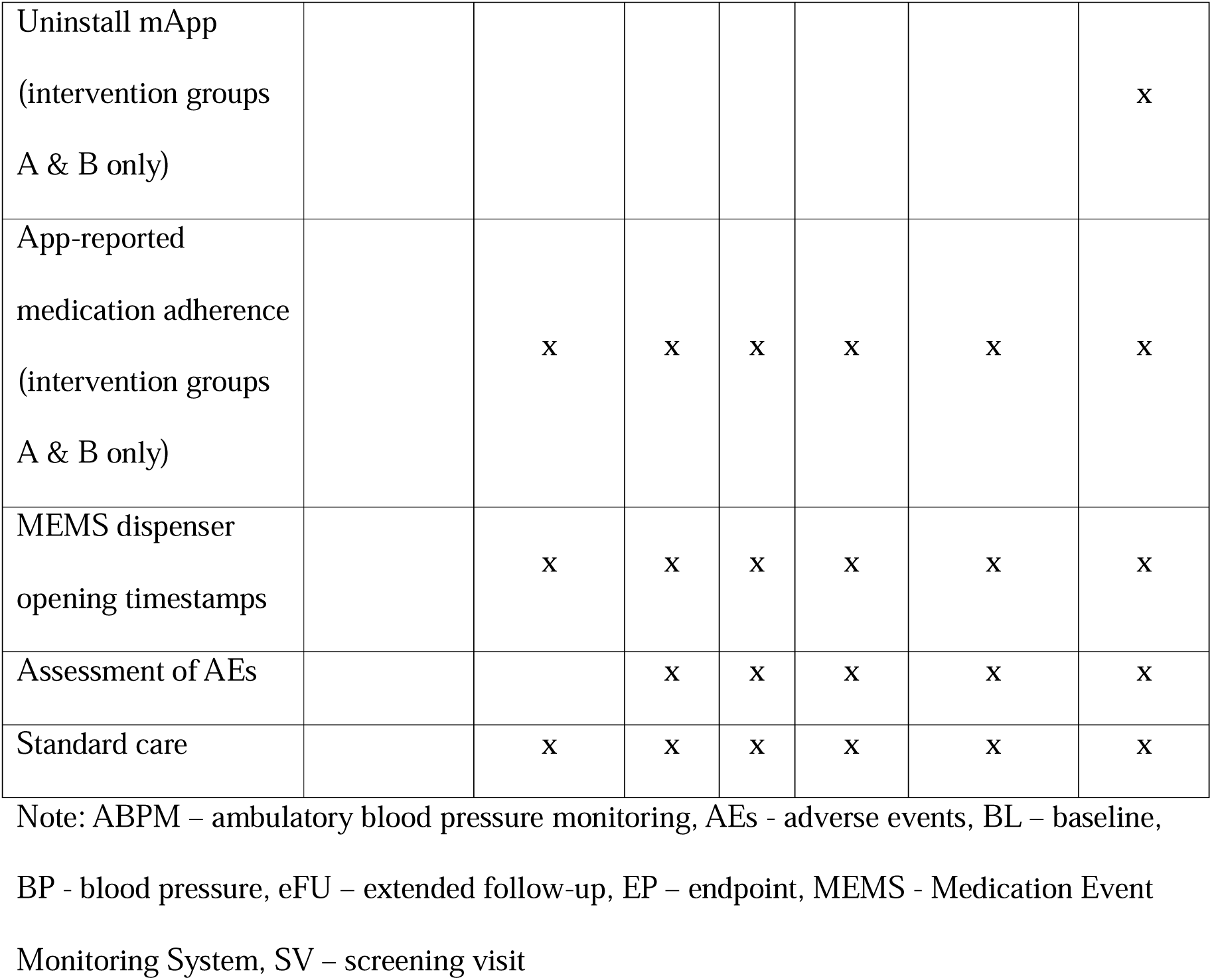
Overview of the study visits, collected data and parameters.

### Participating study centres

The study started as a single-center study at the University Hospital Basel and was expanded to a multi-center study adding four additional sites at University Hospitals in Switzerland (Bern, Zurich, Lausanne, Geneva) due to slow recruitment and the influence of the COVID-19 pandemic.

### Study endpoints and data source

The primary endpoint will be the difference in mean MEMS adherence at EP (90 days) between intervention group A and the control group. The co-primary endpoint will be the MEMS adherence during the last month (Days 61-90) relative to the first month (Days 1-30) of the intervention. The co-primary endpoint was added in an amendment to the initial protocol to account not only for the mean MEMS adherence during the intervention period but also for changes in MEMS adherence over time relative to the individual patients’ starting points (month 3 versus month 1). The secondary endpoints in the trial are listed in Table 3.

**Table 3.**
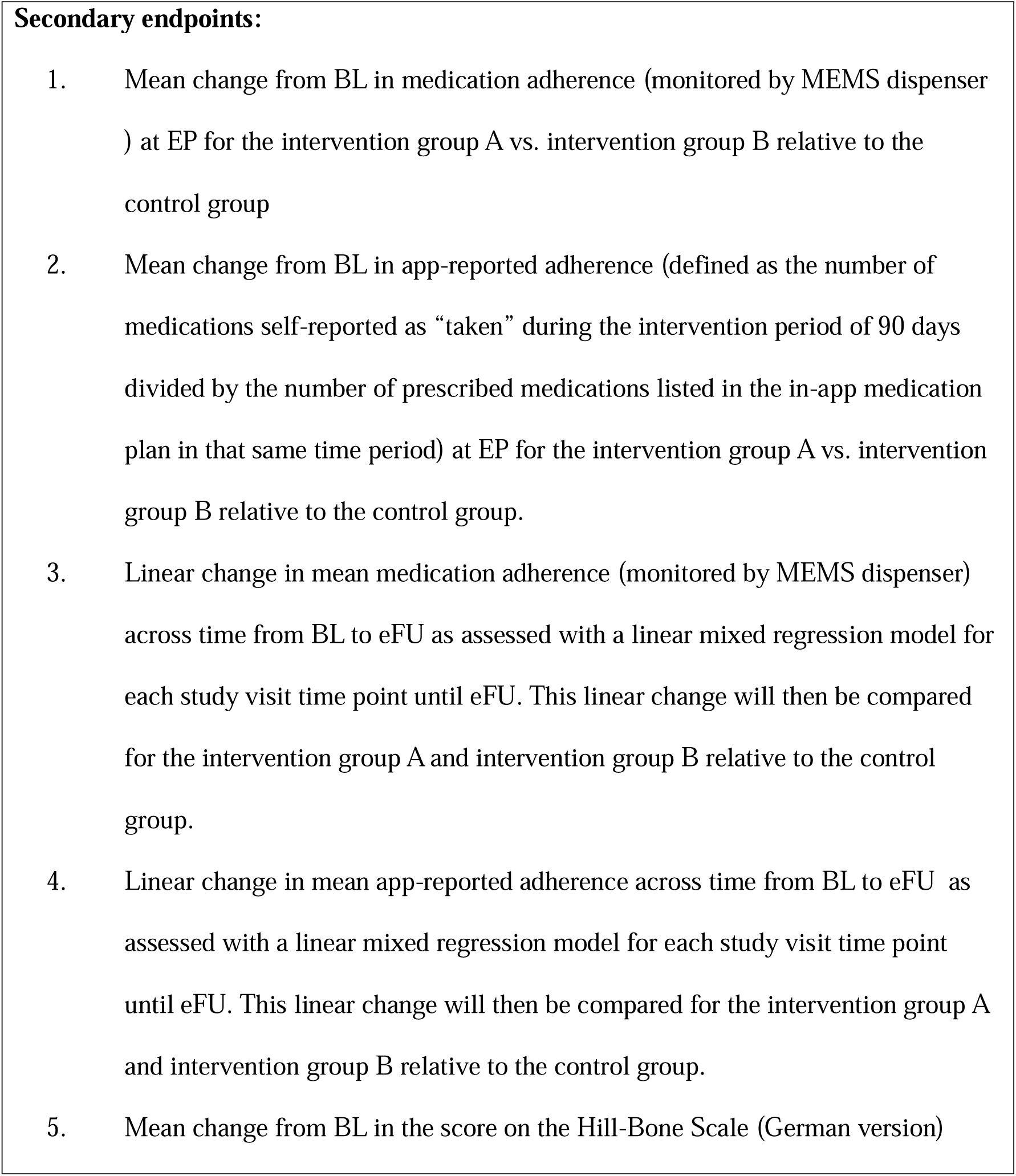
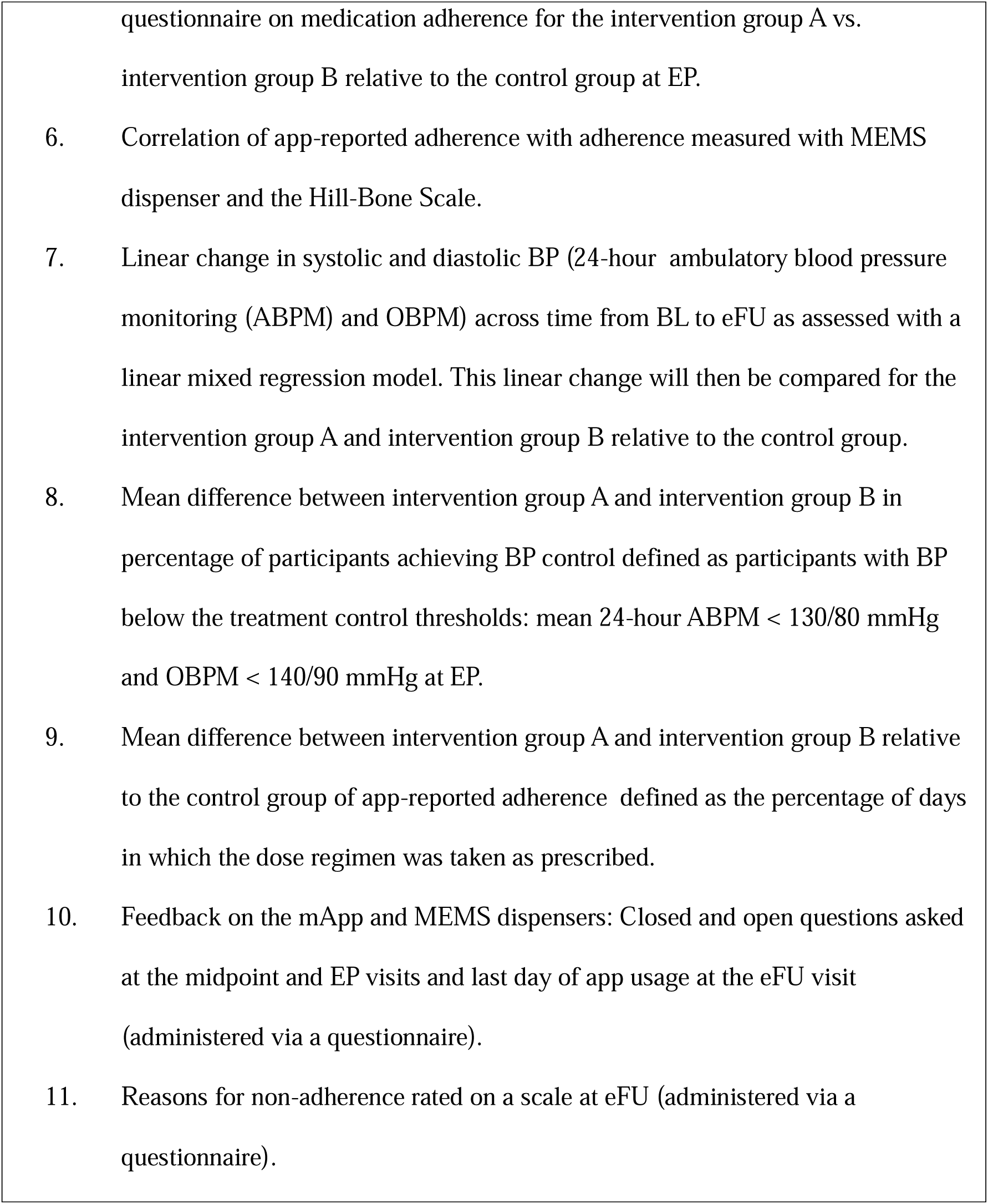
Secondary endpoints.

Participant data will be collected at four in-person visits (BL, 45 days, 90 days and 180 days). Parameters are shown in Table 2. Data will include app-reported adherence, the results of questionnaires, and physical findings such as OBPM and 24-hour ABPM.

### Measurements

All participants will receive MEMS dispensers (Evalan BV, Sarphatistraat 638, 1018 AV Amsterdam, Netherlands) at the BL visits to store their antihypertensive medication. Since participants can store only one medication in the MEMS dispenser, the study team will prioritize the antihypertensive drug to be monitored based on the expected clinical benefit from improved adherence The suggested priority for antihypertensives to be placed in the MEMS dispenser is: 1) any combination preparation 2) Renin-angiotensin system inhibitors, 3) calcium antagonists, 4) diuretics. To determine BP, we use a standard operating procedure based on the European Society of Cardiology/European Society of Hypertension (ESC/ESH) guidelines [2,20]. In brief, BP measures are taken in a seated position after 5 min of rest with feet on floor, back supported; no caffeine, exercise, or smoking during the 30 min before the measurement; emptied bladder; no talking during measurement; comfortable clothes; and arms supported. BP is assessed on the same arm (i.e., the reference arm) at each visit using a validated automated measurement device. At the screening visit (SV), the reference arm is determined by measuring BP on both arms and taking the arm having the higher BP. The BP is calculated as the mean value of the last two out of three consecutive measurements, spaced 1–2 min apart [29].

For the 24-hour ABPM recordings, measurements will always be taken on the non-dominant arm. The device will be programmed to take measurements every 20 minutes between 6:00 and 22:00 (defined as the day-time period) and every 30 minutes between 22:00 and 6:00 (night-time period). 24-hour ABPM is planned at BL, EP, and eFU visits. If a 24-hour ABPM has been taken within 6 weeks prior to the BL visit, the BL 24-hour ABPM may be skipped.

The questionnaires (Hill-Bone Scale Questionnaire, Reasons for Medication Adherence Questionnaire, Prototype Feedback Questionnaires, MEMS dispenser Usability Questionnaire) are administered to the patients after inclusion and completed by the patients in private following the requirements of each questionnaire[14,26].

Participants allocated to the intervention groups will receive the mApp, and the investigator will input the participant’s medications into the mApp. App-reported adherence of medication intake will be carried out by the patients (Figure 2).

**Figure 2.**
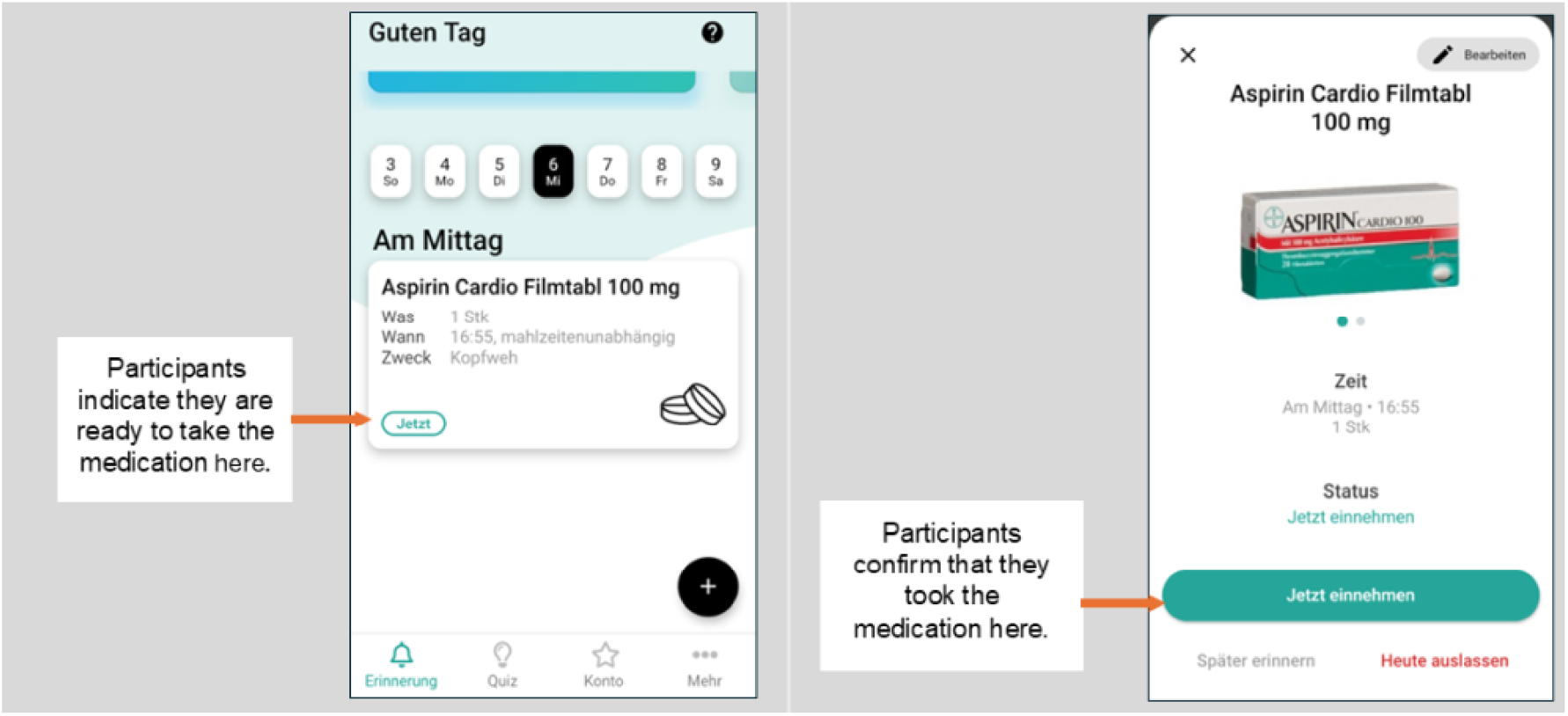
In-mApp self report of medication intake

### Data collection

Data will be collected during personal visits of the participants with the study team at the defined time points in Table 2 and according to the primary and secondary endpoints. At the BL visit, electronic health record will be screened to identify co-morbidities in addition to the personal history taken by the study team. In case of uncertainties family physicians of the participants may be contacted. At each of the following visits, an interim medical history will be taken with a focus on changes of the antihypertensive medication, concomitant medication, adverse events (AEs), and serious adverse events (SAEs). At each visit, continued eligibility will be assessed. In line with a protocol amendment (see Table 1), only changes in antihypertensive medication and not concomitant medication during the intervention phase lead to exclusion.

### Data management

All data collected at the study site will be recorded in an eCRF provided by shareCRF (www.shareCRF.com). For each enrolled study participant, an eCRF will be maintained and kept current to reflect subject status at each phase of the study.

Study-relevant data such as app-reported and objective medication adherence will be collected in the mApp or via the MEMS timestamps. Additional measurements are listed in Table 2. Queries generated by rules and raised by reviewers will be recorded within the electronic data capture (EDC) application. The participants identification numbers (IDs) will be user IDs used to log into the mApp and will be generated ahead of time by the Sponsor. For the control group, participant IDs will be generated similarly but will not be used to as a login. Additionally, and as part of the eCRF, the medications list, AE, and SAE reporting and any withdrawal forms will be treated as source documents. Finally, the data recorded by the mApp are also treated as source documents.

### Statistical considerations

The BEHAVE-HTN study will be evaluated by Full Analysis Set, defined as every randomized participant except those who withdrew or were excluded from the study early due to exclusion criteria, withdrawal of consent, or lack of compliance with the study protocol. Data will be included up to the last visit before exclusion or withdrawal. The null hypothesis for this study is that themApp with financial incentives will not increase medication adherence compared to control group according to the primary endpoints. The alternative hypothesis is that medication adherence will be significantly increased for the intervention group A relative to the control group.

Prior studies showed financial incentives improved medication adherence by 12 percentage points relative to controls in 92 non-adherent glaucoma patients, with an effect size of 0.46 [30]. In order to detect a similar effect, assuming a two-sample t-test, α one-sided = 0.05, and power of 0.8, we would need to recruit a sample of 60 participants in each group, with 180 participants in total. We will employ a sequential hypothesis testing procedure using Bayes Factors (BFs) to assess the relative strength of evidence for the alternative compared to the null hypothesis at certain time points [31,32]. We plan to conduct an initial assessment after 20 participants as the recommended minimum sample size for sequential hypothesis testing have completed the study in each group[31]. At this point we will begin monitoring the BF. If we achieve either a BF >= 6 or a BF <=1/6, which would indicate substantial evidence for the alternative or null hypotheses, respectively, we will stop the study [33–35]. Otherwise, we will adjust the sample size until we reach either a BF ≥ 6 (supporting the alternative hypothesis), a BF ≤ 1/6 (supporting the null hypothesis) or reach the final sample size of 180 participants.

The primary and co-primary endpoints will be analyzed using a two-sample Bayesian t-test [35]. A posterior probability > 0.95 in favor of the alternative hypothesis will be considered significant, and a BF > 6 supporting the alternative hypothesis will be considered substantial evidence of an effect [33–35]. The secondary endpoints 1-5 and 7-9 will be analyzed using Bayesian regression models [36]. Secondary endpoint 6 will be analyzed using a Bayesian Pearson’s correlation. As the study is not stratified by any other variables other than gender, we will conduct additional analyses with the covariates of participant age as well as number of pills per day taken by the participants. The primary and co-primary endpoints as well as secondary outcome measures will be analyzed using multiple regression including participant age and number of pills taken included as a covariate, and the answers for secondary endpoints 10 and 11 will be coded with a Likert scale and will be described with a summary statistic.

Missing data points on medication intake from themApp as well as the MEMS dispenser will be classified as participants being non-adherent.

Data analysis will be performed using the statistical tools available in IBM SPSS Statistics and R [37,38]. Data will remain available for later secondary analysis for up to 10 years.

### Consent and Ethics

The study complies with local legal requirements and has been approved by the ethics committee northwestern and central Switzerland (2021-00279). The study will be carried out in accordance with the protocol and with principles enunciated in the current version of the Declaration of Helsinki, and all patients must provide written informed consent to participate.

### Registration

The study is registered at SNCTP - Swiss National Clinical Trials Portal (SNCTP000004345) and at ClinicalTrials.gov (NCT04708756).

## Results

Currently, participant inclusion has been nearly completed. We are undergoing data clearing and are planning data analysis. These steps are planned for autumn 2025.

## Discussion

Low patient adherence remains one of the key factors of suboptimal BP control in the management of hypertension [2]. Consequences of low adherence are wide-ranging and include not only a higher risk for morbidity and mortality for the patients but also a large increase in costs for the healthcare system and a negative influence on sustainability through redeemed but unused prescriptions [39].

Given a prevalence of patient adherence to regular drug intake at only 30–50 %, any effective approaches to improving adherence are critical [40] and improving adherence of patients with pharmacological therapy is a highly potent therapeutic approach to decelerate disease progression and improve in prognosis. For this reason, the 2023 ESH guidelines put a particularly strong emphasis on the detection and management of non-adherence to BP-lowering therapy [41].

According the ESC/ESH guidelines the management of non-adherence to antihypertensive treatment should be tailored to the individual modifiable drivers of non-adherence in each patient [41]. There is not a single universal strategy that could help to manage non-adherence in all hypertensive patients.

The use of mobile-based technologies to support health care is on the rise owing to their easy implementation, low cost, accessibility across time and location, and minimizing the issue of distance in access to services [42]. These advantages facilitated the use of mobile-based applications as potential tools to improve medication adherence. Currently, there is a plethora of available mobile apps aimed at improving medication adherence; however, there is little evidence evaluating these interventions for efficacy. In a study by Bakes et al executed in Switzerland, the authors identified 1833 free-of-charge applications and 307 paid applications and after a rigorous eligibility check only included 4 vs. 3 mHealth applications for further evaluation regarding security and privacy, quality of the health-related content, quality of the app information management, functionality, user interface and acceptability of the app. They conclude that, to date, healthcare providers should be discouraged from recommending these applications[43]. On the other hand, a systematic review and meta-analysis in the American Journal of Preventive Medicine found significant improvements in medication adherence among patients with hypertension by the use of mHealth applications[22].

The multi-center, prospective, and randomized BEHAVE-HTN study will investigate whether a multi-faceted medication adherence mobile application based intervention including reminders, information, gamification, and financial incentives will improve adherence compared to standard of care. Furthermore, by adding an intervention group without financial incentives, it will help to distinguish between the effect of the mApp and that of financial incentives. The two phases of the study – an initial intervention phase (day 0-90) and a follow-up phase (day 90-180) – will also enable us to investigate whether sustainable routines can be formed beyond the intervention and whether these also have an influence on further endpoints such as BP control. The planned study provides a rigorous design, a sufficient size of participants, and covers three different regimens. The strengths of the study are the differentiated primary endpoints that contain an evaluation of several highly relevant behavioural changes.

There are possible limitations of the proposed approach. First, the control group will receive standard care only and will not receive the mApp or any other intervention. Second, despite existing evidence that 90 days is an adequate time to establish sustained behavioral changes, we are unable to exclude that long-term effects of the intervention may vanish over the time after trial conclusion [44]. Electronic drug monitoring using sensors that register an act of opening a medication-dispensing container/blister pack is generally very accurate and provides detailed information of both the timing and frequency of adherence as well as persistence on treatment [41]. However, the costs, the risk of intentional/unintentional dose removal from the container without ingestion, and the infeasibility of the method for monitoring numerous antihypertensive medications also represent important limitations.

After the initiation of the study, we faced several difficulties in the execution of the trial leading to adaptions of the study protocol. One major aspect was the COVID-19 pandemic, which had a significant impact on the execution of clinical studies. Government measures had significant consequences on healthcare, particularly in the management of chronic diseases such as hypertension. Many patients, especially those considered more vulnerable due to age, immunosuppression, or other factors, began to avoid hospitals and healthcare facilities out of fear of infection. On the other hand, more resources were allocated to combating the COVID-19 pandemic, leading to a reprioritization of which clinical studies were conducted. This reluctance also affected outpatient treatment, including at the Medical Outpatient Department in Basel.

There was compromised medical care for hypertensive patients during the COVID-19-related shutdowns that affected medical consultations, diagnostic measures, BP monitoring, and questions regarding the medication intake and may have led to the discontinuation of the medication regimen[45,46]. Additionally, concerns arose regarding certain antihypertensive drugs, specifically Renin-Angiotensin System inhibitors, which were being investigated for their potential connection to the risk and severity of COVID-19 infection. These concerns led to the discontinuation of such therapies and changes in antihypertensive treatment schedules[47]. For the BEHAVE-HTN study, we faced a slower than expected recruitment rate and a higher than expected, dropout rate due to changes in concomitant medications, withdrawals of participants, missed consultations, and delayed payments of incentives. To account for these influences and difficulties we decided to only exclude patients if changes in antihypertensive medication occurred during the intervention phase and not in case of changes in concomitant medication and to expand the trial to four additional sites at tertiary centres in Switzerland.

Taken together, the results of the BEHAVE HTN study will offer valuable insights into the effectiveness of the mApp, both with and without financial incentives, and will assess the value of a behavioural economics-based approach, in managing BP, while also identifying barriers to medication adherence.

## Data Availability

This is a protocol publication. Further published data will be available upon reasonable request to the authors

## Acknowledgements

We thank Laurie Theurer serving as external monitor for the study.

## Data availability

All data are made available by the corresponding author upon reasonable request.

## Autor contributions

**Conceptualization:** ARB, ASV, TB**, Formal analysis:** ASV, JS**, Funding acquisition:** ARB**, Investigation:** ASV, JS, GW, GE, MS, ER, TM, TB**, Methodology:** ARB, MM, ASV, TB; **Project administration:** ASV, ARB, GW, GE, MS, ER, TM, TB, LT **; Resources:** ARB, MM, GW, GE, MS, ER, TM, TB**; Supervision:** ASV, ARB, GW, GE, MS, ER, TM, TB, LT**; Writing original draft:** ASV, JS, VN, TB**; Writing – review & editing:** ARB, ASV, JS, VN MM, GW, GE, MS, ER, TM, TB

## Abbreviations

ABPM: Ambulatory blood pressure monitoring
AE(s): Adverse event(s)
BF(s): Bayes Factor(s)
BL: Baseline
BP: Blood pressure
eCRF: Electronic Case Report Form
IDs: Identification numbers
mApp: Mobile application
MEMS: Medication Event Monitoring System
OBPM: Office blood pressure monitoring
SAE(s): Serious adverse event(s)
SV: Screening visit
WHO: World Health Organization

